# Predicting polycystic ovary syndrome (PCOS) with machine learning algorithms from electronic health records

**DOI:** 10.1101/2023.07.27.23293255

**Authors:** Zahra Zad, Victoria S. Jiang, Amber T. Wolf, Taiyao Wang, J. Jojo Cheng, Ioannis Ch. Paschalidis, Shruthi Mahalingaiah

**Affiliations:** Division of Systems Engineering, Center for Information and Systems Engineering (CISE), Boston University, 15 St. Mary’s Street, Brookline, MA 02446, USA; Division of Reproductive Endocrinology and Infertility, Department of Obstetrics and Gynecology, Massachusetts General Hospital, 55 Fruit Street, Yawkey 10, Boston, MA 02114, USA; Icahn School of Medicine at Mount Sinai, 1 Gustave L. Levy Place, New York, NY 10029, USA; Department of Biostatistics and Medical Informatics, University of Wisconsin, West Johnson Street, Madison, WI 53792, USA; Department of Electrical & Computer Engineering, Department of Biomedical Engineering, and Faculty for Computing & Data Sciences, Boston University, 8 St. Mary’s Street, Boston, MA 02215, USA; Department of Environmental Health, Harvard T.H. Chan School of Public Health, 665 Huntington Avenue, Boston, MA 02115, USA

**Keywords:** Polycystic ovary syndrome (PCOS), disease prediction, predictive model, machine learning, artificial intelligence

## Abstract

**Introduction:** Predictive models have been used to aid early diagnosis of PCOS, though existing models are based on small sample sizes and limited to fertility clinic populations. We built a predictive model using machine learning algorithms based on an outpatient population at risk for PCOS to predict risk and facilitate earlier diagnosis, particularly among those who meet diagnostic criteria but have not received a diagnosis.

**Methods:** This is a retrospective cohort study from a SafetyNet hospital’s electronic health records (EHR) from 2003-2016. The study population included 30,601 women aged 18-45 years without concurrent endocrinopathy who had any visit to Boston Medical Center for primary care, obstetrics and gynecology, endocrinology, family medicine, or general internal medicine. Four prediction outcomes were assessed for PCOS. The first outcome was PCOS ICD-9 diagnosis with additional model outcomes of algorithm-defined PCOS. The latter was based on Rotterdam criteria and merging laboratory values, radiographic imaging, and ICD data from the EHR to define irregular menstruation, hyperandrogenism, and polycystic ovarian morphology on ultrasound.

**Results:** We developed predictive models using four machine learning methods: logistic regression, supported vector machine, gradient boosted trees, and random forests. Hormone values (follicle-stimulating hormone, luteinizing hormone, estradiol, and sex hormone binding globulin) were combined to create a multilayer perceptron score using a neural network classifier. Prediction of PCOS prior to clinical diagnosis in an out-of-sample test set of patients achieved AUC of 85%, 81%, 80%, and 82%, respectively in Models I, II, III and IV. Significant positive predictors of PCOS diagnosis across models included hormone levels and obesity; negative predictors included gravidity and positive bHCG.

**Conclusions:** Machine learning algorithms were used to predict PCOS based on a large at-risk population. This approach may guide early detection of PCOS within EHR-interfaced populations to facilitate counseling and interventions that may reduce long-term health consequences. Our model illustrates the potential benefits of an artificial intelligence-enabled provider assistance tool that can be integrated into the EHR to reduce delays in diagnosis. However, model validation in other hospital-based populations is necessary.

## Introduction

Polycystic ovary syndrome (PCOS) is the most common type of ovulation disorder and endocrinopathy among reproductive age women. PCOS is a diagnosis of exclusion after other endocrinopathies known to affect ovulation have been evaluated including thyroid, adrenal, and pituitary related disease. Based on the Rotterdam criteria, PCOS is diagnosed when two of the three following criteria are exhibited: clinical or biochemical hyperandrogenism, oligo-anovulation, and polycystic ovary morphology (PCOM) on transvaginal or transabdominal ultrasound. PCOS has a population prevalence of 5-15%, depending on the diagnostic criteria used (1).

PCOS is associated with multiple health issues and increased morbidity and mortality, including a high chronic disease burden that is also very costly for individuals with PCOS and insurers (2). PCOS is the leading cause of anovulatory infertility in reproductive-aged women. In fact, over 90% of anovulatory women who present to infertility clinics have PCOS (3). PCOS patients have an increased risk of endometrial hyperplasia and endometrial cancer (4) due to anovulatory cycles leading to long periods of exposure to the effects of unopposed estrogen. PCOS has been associated with the development of metabolic syndrome (5), diabetes (6), cerebrovascular disease and hypertension (7), compared to women without PCOS. Despite these serious health consequences, PCOS frequently goes undiagnosed due to the wide range of symptom severity on presentation, leading to delayed treatment and potentially more severe clinical sequelae due to lack of preventive care, health management, and counseling (4). Even when PCOS is diagnosed, it is often very delayed. One study found that over one-third of women with PCOS waited over two years and were seen by three or more providers before finally receiving the diagnosis (8).

Predictive models can play a significant role in aiding earlier diagnosis of PCOS, though several include only those women presenting for fertility care. One model used serum anti-Müllerian hormone (AMH) and androstenedione levels, menstrual cycle length, and BMI to predict the development of PCOS in Chinese women (9). Another model used only AMH and BMI to predict a diagnosis of PCOS or other ovulatory dysfunction disorders (10). Other studies have created predictive models for certain outcomes among women with PCOS such as pregnancy outcomes (11,12) and insulin resistance (13). In this study, we use clinical and socioeconomic variables among 30,601 women aged 18 to 45 years within the electronic health records (EHR) to develop predictive model utilizing machine learning algorithms with the goal of earlier detection and treatment of PCOS.

## Materials and Methods

### Data acquisition

The dataset was created by querying de-identified patient data from female patients aged 18 to 45 years who had or were considered at risk for PCOS diagnosis by having had any one of the three testing procedures for PCOS in their EHR. Included within the initial sample were those patients who had any visit to Boston Medical Center (BMC) for primary care, obstetrics and gynecology, endocrinology, family medicine, or general internal medicine and received: 1) a pelvic/transvaginal ultrasound for any reason, 2) androgen lab assessment, or had clinical symptoms of androgen excess, 3) an ICD-9 label for irregular periods, or 4) a PCOS diagnosis, between October 2003 to December 2016 within the BMC Clinical Data Warehouse (CDW). The start-date was selected to reflect the first day that ICD-9 codes were used and recorded at BMC. The end date reflected cessation of use of the ICD-9 codes and transition to ICD-10 codes within BMC. To avoid misidentifying an ovulation disorder caused by another endocrinopathy, exclusion criteria included diagnosis of concurrent endocrinopathy, such as thyroid disorders, hyperaldosteronism, Cushing’s syndrome, other adrenal gland disorders, or malignancy based on ICD-9 codes as listed in Supplementary Table 1.

### Ethical approval

The study was approved by the Institutional Review Board of Boston University School of Medicine and the Harvard T.H. Chan School of Public Health (Protocol # H35708) and is considered non-human subjects research.

### Reference label definitions

#### Individual predictors

Time-varying predictor variables with a date stamp before that of the outcome of interest were included in our models. We considered the following predictor variables:

##### Socioeconomic and lifestyle demographic variables

age, race (White/Caucasian, Black/African American, Hispanic/Latina, Asian, Native Hawaiian/Pacific Islander, Middle Eastern, Other/Unknown), smoking status (yes/no), marital status (single, married, separated, divorced, widowed, other), homelessness (yes/no), and highest level of education (8^th^ grade or less, some high school, high school graduate, some college/technical/vocational training, graduated college/technical school/vocational training, declined to answer, other).

##### Anthropometrics

Body mass index (BMI, kg/m^2^) was either calculated from height and weight or abstracted as the listed BMI variable associated with each visit. BMI was then categorized into three categories: normal (BMI < 25 kg/m^2^); overweight (BMI between 25-30 kg/m^2^); and obese (BMI > 30 kg/m^2^). To further capture the obesity population in the absence of height/weight/BMI data, the obese category also included any patient with an ICD-9 code for unspecified obesity (278.00), morbid obesity (278.01), localized adiposity (278.1), and/or a history of gastric bypass.

##### Cardiovascular health

To include blood pressure as a predictor variable, we defined a categorical hypertension variable by using systolic (SBP) and diastolic (DBP) blood pressure readings and ICD-9 diagnostic codes for unspecified essential hypertension (401.9), benign essential hypertension (401.1), and essential primary hypertension (401.0). Blood pressure was categorized into three groups: normal, defined by no ICD-9 codes for hypertension recorded and SBP < 120 mmHg, and DBP < 80 mmHg; elevated, defined by no ICD-9 codes for hypertension recorded and SBP was 120-129 mmHg or DBP < 80 mmHg; hypertension, defined by any ICD-9 code for hypertension recorded or SBP ≥ 140 mmHg or DBP ≥ 90 mmHg.

##### Reproductive endocrine predictive variables

beta human chorionic gonadotropin (bHCG) level (negative bHCG < 5 mIU/mL, positive bHCG ≥ 5 mIU/mL), HIV status (negative/positive), age at menarche, pelvic inflammatory disease diagnosis (614.9), history of hysterosalpingogram, and gravidity (history of present or prior pregnancy within obstetric history). Endocrine and metabolic lab values included: TSH, glycosylated hemoglobin (A1c) as a marker for diabetes, low-density lipoprotein (LDL), high density lipoprotein (HDL), and diagnosis of hypercholesterolemia (272.0). Of note, our model did not include androgen precursors such as DHEA or androstenedione as, according to Monash guidelines, these values provide limited additional information in the diagnosis of PCOS (14,15).

### Combined predictors

Expecting a nonlinear relationship between many reproductive hormones and a PCOS diagnosis, we used a multilayer perceptron (MLP) neural network to map follicle-stimulating hormone (FSH), luteinizing hormone (LH), sex hormone binding globulin (SHBG), and estradiol (E2) values to a composite metric we call MLP score. The MLP score was repetitively trained and the hyperparameters were tuned to generate a predictive probability associated with PCOS diagnosis for each predictive model, as described with further detail below.

### Outcomes

#### Defining PCOS

PCOS diagnosis was assigned for any patient who had an ICD-9 code for PCOS (256.4) or met the Rotterdam criteria (16), according to which a positive diagnosis is made in the presence of two out of the following three features: (i) irregular menses (IM) as defined by rare menses, oligo-ovulation, or anovulation; (ii) hyperandrogenism (HA) as defined by clinical or biochemical androgen excess; and (iii) polycystic ovarian morphology (PCOM) noted on transabdominal or transvaginal ultrasound. Based on these three criteria, we defined three auxiliary variables IM, HA, and PCOM to use in the definition of our labels. PCOM was captured through diagnostic radiology text reports from ovarian ultrasound imaging for the subset that had ultrasound imaging (17).

#### Defining Irregular Menstruation (IM)

IM was defined with the following ICD-9 codes: absence of menstruation (626.0), scanty or infrequent menstruation (626.1), irregular menstrual cycle (626.4), unspecified disorders of menstruation and abnormal bleeding from female genital tract (626.9), and infertility, female associated with anovulation (628.0) (3).

#### Defining Hyperandrogenism (HA)

HA was assigned to a patient if any of the androgen lab testing for bioavailable testosterone, free testosterone, or total testosterone was greater than clinical thresholds of 11 ng/dL, 5 pg/mL, 45 ng/dL, respectively. In addition, HA was assigned if ICD-9 codes for hirsutism (704.1) or acne (706.1 or 706.0) were recorded for a patient.

#### Defining Ultrasound characteristics for polycystic ovarian morphology (PCOM)

Among those with an ultrasound in this dataset, PCOM was identified on ultrasound reports using natural language processing (NLP) with complete methods detailed by Cheng and Mahalingaiah (17), to report PCOM as identified (PCOM present), unidentified (PCOM absent), or indeterminate (PCOM unidentifiable based on source report data).

We considered four models to predict the following: **Model I**: patients with ICD-9 diagnosis of PCOS (256.4) within the EHR; **Model II**: patients diagnosed with PCOS by Rotterdam criteria having IM and HA without a specific ICD-9 PCOS code; **Model III**: patients diagnosed with PCOS by Rotterdam criteria having two out of the three conditions IM/HA/PCOM and without a specific ICD-9 PCOS code; **Model IV**: all patients with PCOS using either Model I or Model III criteria. ICD-9 codes were abstracted from the billing code and diagnosis code associated with each encounter within the EHR. Model I included all patients who were diagnosed with PCOS. Model II and its superset Model III was composed of patients who did not have a PCOS diagnosis code but met diagnostic criteria of PCOS based on Rotterdam criteria, representing the patient population with undiagnosed PCOS. Model IV essentially captures all women who were diagnosed or met criteria for PCOS within our population. Supplementary Table 2 details model definitions and includes the count and percent of patients in each category. The date of diagnosis was assigned by the date of PCOS ICD-9 code (256.4) for Model I, the date of the latest diagnostic criteria met for Model II and III, and the earlier date associated with Model I and Model III, for Model IV.

### Predictive models

#### Classification methods

We explored a variety of supervised classification methods, both linear and nonlinear. Linear methods included logistic regression (LR) and support vector machines (SVM) (18) and were fitted with an additional regularization term: an L1-norm of the coefficient vector to inject robustness (19) and induce sparsity. Regularization added a penalty to the objective function, thereby minimizing the sum of a metric capturing fitness to the data and a penalty term that is equal to some multiple of a norm of the model parameters. Sparsity was motivated by the earlier works (20–23), where it was shown that sparse classifiers can perform almost as well as very sophisticated classification methods. Nonlinear methods, including gradient boosted trees (GBT/XGBoost) (24) and random forests (RF) (25) which produce large ensembles of decision trees, may yield better classification performance, but are not interpretable or explainable to enable a safety check by a clinician. Specifically, the RF is a large collection of decision trees and it classifies by averaging the decisions of these trees. The GBT/XGBoost, also called gradient boosting machine (GBM), similarly combines decisions by many decision trees. We used LightGBM which is a fast, high-performance GBM framework (26). We tuned GBM’s hyperparameters through cross-validation.

#### Performance metrics

To assess model performance, we obtained the Receiver Operating Characteristic (ROC) curve. The ROC is created by plotting the true positive rate, which is indicative of sensitivity or recall, against the false positive rate (equal to one minus specificity) at various thresholds. The c-statistic or the area under the ROC curve (AUC), is used to evaluate the prediction performance. A perfect predictor is defined by generating an AUC score of 1, and a predictor which makes random guesses has an AUC score of 0.5. We also used the weighted-F1 score to evaluate the models. The weighted-F1 score is the average of the F1 scores of each class weighted by the number of participants in each class. The class-specific F1 scores are computed as the harmonic mean of precision and recall of a classifier which predicts the label of the given class. The weighted-F1 score is between 0 to 1, and a higher value represents a better model. The AUC is more easily interpretable, and the weighted F1-score is more robust to class imbalance (27).

#### Statistical feature selection (SFS)

Categorical variables were converted into dummy/indicator variables. To avoid collinearity, we dropped the missing or unclassified data (NaN) category. For continuous variables, missing values were imputed by the median value for that variable. A summary of the missing variables for each model is provided in Supplementary Table 3. Variables with very low variability (SD<0.0001) were assessed for removal from the models, however none were noted in any model. We applied statistical feature selection (SFS) to reduce the less informative features and simplify the models. For each of the four models’ outcomes, the chi-squared test was applied for binary variables and the Kolmogorov-Smirnov statistic for continuous variables; the variables for which we could not reject the null hypothesis of the same distribution for each class (p-value >0.01) were removed. Representative aggregated patient-level statistics for each model are shown in Supplementary Table 4. We also removed one from each pair of highly correlated variables (with absolute value of the correlation coefficient > 0.8) to avoid redundant variables. Highly correlated variables and the retained variable are provided in Supplementary Table 5. For all models we standardized the corresponding features by subtracting the mean and scaling to unit variance.

#### Training-test splitting

We split the dataset into five random parts, where four parts were used as the training set, and the remaining part was used for testing. We used the training set to tune the model hyperparameters via 5-fold cross-validation, and we evaluated the performance metrics on the testing set. We repeated training and testing five times, each time with a different random split into training/test sets. The mean and standard deviation of the metrics on the test sets over the five repetitions are reported.

#### Development of the MLP score

For every model, there was a considerable difference between the AUC of linear models and non-linear models. To improve the performance of our linear models, we utilized nonlinear models to capture intricate relationships between features. We utilized Gradient Boosted Trees (GBT) to find which features most commonly appeared together among decision trees. We found FSH, LH, SHBG, and estradiol levels to be a meaningful group of features which are all reproductive hormones and continuous variables that appeared together among trees for all our models. We subsequently used these four features as input features into a multilayer perceptron (MLP) neural network model with three hidden layers, each employing the rectified linear unit (ReLU) activation function. The neural network was trained using the training set to classify PCOS. We used the output probability of the MLP model, which we called “MLP score,” as a new feature into our original predictive models.

#### Recursive feature elimination (RFE)

We also used a recursive feature elimination approach with L1-penalized logistic regression (L1-regularized RFE) to extract the most informative features and develop parsimonious models. Specifically, after running the L1-penalized logistic regression (L1-LR), we obtained weights associated with the variables (i.e., the coefficients of the model, denoted by β), and we eliminated the variable with the smallest absolute weight in each turn. We iterated in this fashion, eliminating one variable at a time, to select a model that maximizes a metric equal to the mean AUC minus the standard deviation (SD) of the AUC in a validation dataset (using 5-fold cross-validation on the training set to obtain an average of this metric over five repetitions).

#### Final predictive models

We computed the performance of the following models: L1-penalized logistic regression (LR-L1), support vector machine (SVM-L1), random forests (RF), and gradient boosted trees (GBT/XGBoost). We calculated each variable’s LR coefficient with a 95% confidence interval (β [95%CI]), the correlation of the variable with the outcome (Y-correlation), the p-value of each variable (p-value), the mean of the variable (Y1-mean) in the PCOS labeled patients, the mean of the variable (Y0-mean) in the patients without the PCOS label, and the mean and standard deviation of the variable over all patients (All-mean and All-SD). Ranking predictor variables by the absolute value of their coefficients in the logistic regression model amounts to ranking these variables by how much they affect the predicted probability of the outcome. A positive coefficient implies that the larger the value of the variable within the range specified by the data, the higher the chance of having a PCOS diagnosis as defined by the model outcome.

## Results

### Results of data acquisition and data pre-processing

After inclusion and exclusion criteria were applied to all 65,431 women within the initial data pool, 30,601 patient records were available for this analysis and defined populations are included in Figure 1. There were 1,329 patients (4.5%) with a PCOS ICD-9 diagnosis code (Model I). 1,465 patients had records with PCOM results as present, absent, or unidentifiable. There were 1,056 patients (3.6%) with undiagnosed PCOS (Model II), and a total of 1,116 (3.8%) of patients with no ICD 256.4 indication and two out of IM/HA/PCOM positive criteria (Model III). Finally, there were 2,445 PCOS patients (8.0%) in the combined analysis (Model IV). The total number of records in each model are included in Supplementary Table 2. In the total cohort, the patients were predominantly Black/African American (40.3%) and White (26.5%), with an average age of 33.6 years (SD = 6.6). Complete demographic characteristics are described in Table 1.

**Figure 1.**
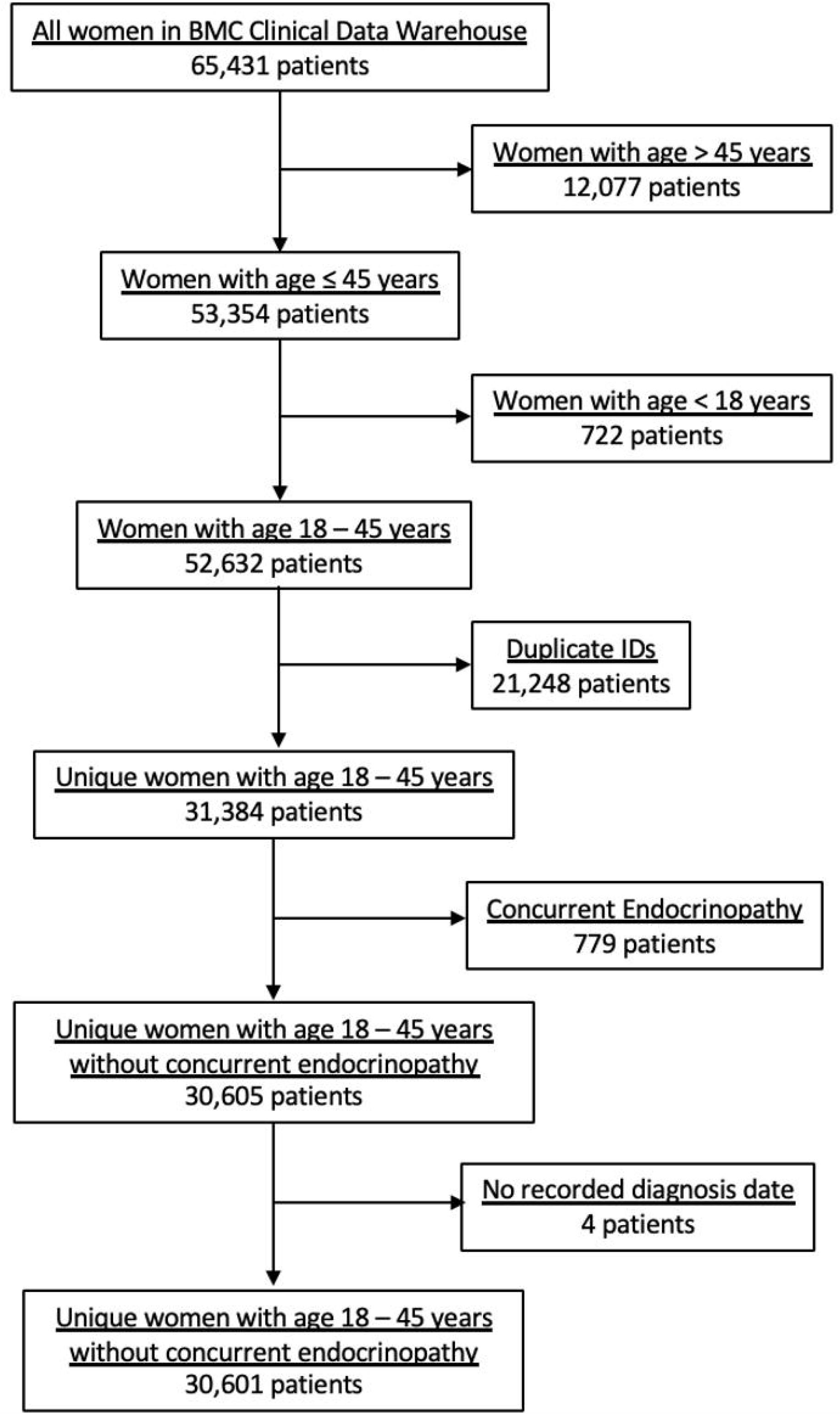
Flow of patients from the BMC CDW into the dataset used by the study.

**Table 1.** Demographic characteristics of the study population and by model.

| Variable | Model I | Model II | Model III | Model IV |
| --- | --- | --- | --- | --- |
| Age, Mean years (SD) | 33.6 (6.6) | 33.7 (6.6) | 33.7 (6.6) | 33.6 (6.6) |
| **Race, n (%)** |  |  |  |  |
| Black/African American | 11881 (40.3) | 11824 (40.5) | 11861 (40.5) | 12395 (40.5) |
| White/Caucasian | 7812 (26.5) | 7733 (26.5) | 7741 (26.4) | 8086 (26.4) |
| Hispanic/Latina | 2858 (9.7) | 2837 (9.7) | 2841 (9.7) | 2929 (9.6) |
| Asian | 1350 (4.6) | 1354 (4.6) | 1354 (4.6) | 1406 (4.6) |
| Middle Eastern | 175 (0.6) | 176 (0.6) | 176 (0.6) | 184 (0.6) |
| American Indian/Native American | 163 (0.6) | 162 (0.6) | 162 (0.6) | 168 (0.5) |
| Native Hawaiian/Pacific Islander | 17 (0.1) | 18 (0.1) | 18 (0.1) | 18 (0.1) |
| Other | 979 (3.3) | 966 (3.3) | 966 (3.3) | 1023 (3.3) |
| Unknown | 4250 (14.41) | 4146 (14.19) | 4153 (14.19) | 4392 (14.4) |
| **Marital Status** |  |  |  |  |
| Single | 22325 (75.7) | 22155 (75.8) | 22199 (75.8) | 23224 (75.9) |
| Married | 5833 (19.8) | 5753 (19.7) | 5767 (19.7) | 6018 (19.7) |
| Separated | 392 (1.3) | 391 (1.3) | 392 (1.3) | 401 (1.3) |
| Divorced | 388 (1.3) | 379 (1.3) | 380 (1.3) | 397 (1.3) |
| Widowed | 35 (0.1) | 35 (0.1) | 35 (0.1) | 35 (0.1) |
| Other | 502 (1.7) | 489 (1.7) | 489 (1.7) | 516 (1.7) |
| Unknown | 10 (0.03) | 10 (0.03) | 10 (0.03) | 10 (0.03) |
| **Body Mass Index** |  |  |  |  |
| Normal (BMI < 25) | 7534 (25.6) | 7685 (26.3) | 7697 (26.3) | 7902 (25.8) |
| Overweight (BMI between 25-30) | 5694 (19.3) | 5689 (19.5) | 5707 (19.5) | 5941 (19.4) |
| Obese (BMI ≥ 30) | 7645 (25.9) | 7369 (25.2) | 7387 (25.2) | 7985 (26.1) |
| Unknown | 8612 (29.2) | 8469 (29.0) | 8481 (29.0) | 8,773 (28.7) |

There were 43 categorical variables and 12 continuous variables retained as predictors after the data pre-processing procedures. There were four pairs of highly correlated variables and one variable from each correlated pair included in the final model as noted in Supplemental Table 5. Supplementary Table 4 describes all 51 variables used by the predictive models.

### Model Performance

Tables 2, 3, 4 and 5 display the parsimonious models that use the MLP score (LR-L2-MLP score) and show the most significant variables in the prediction of the outcome for Models I, II, III, and IV, respectively. All p-values were less than 0.05, which was set as the significance level.

**Table 2.**
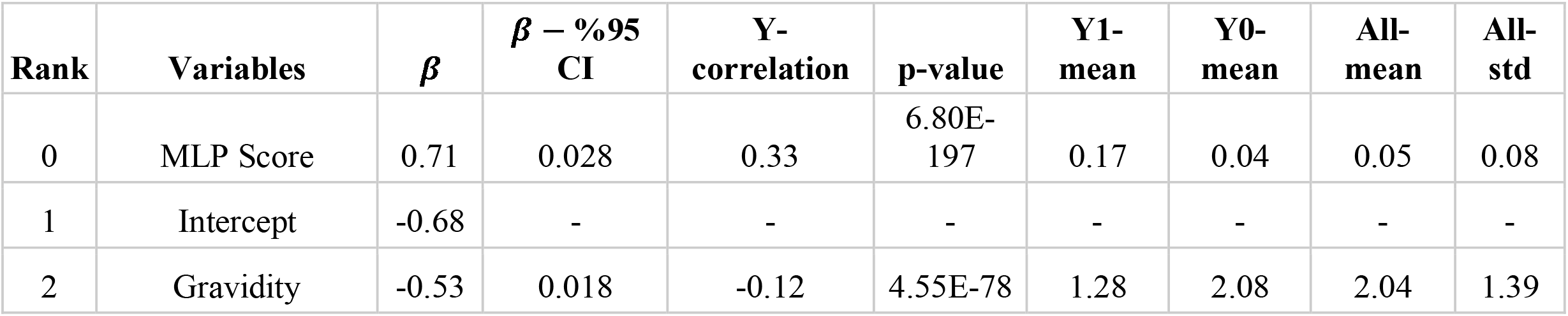
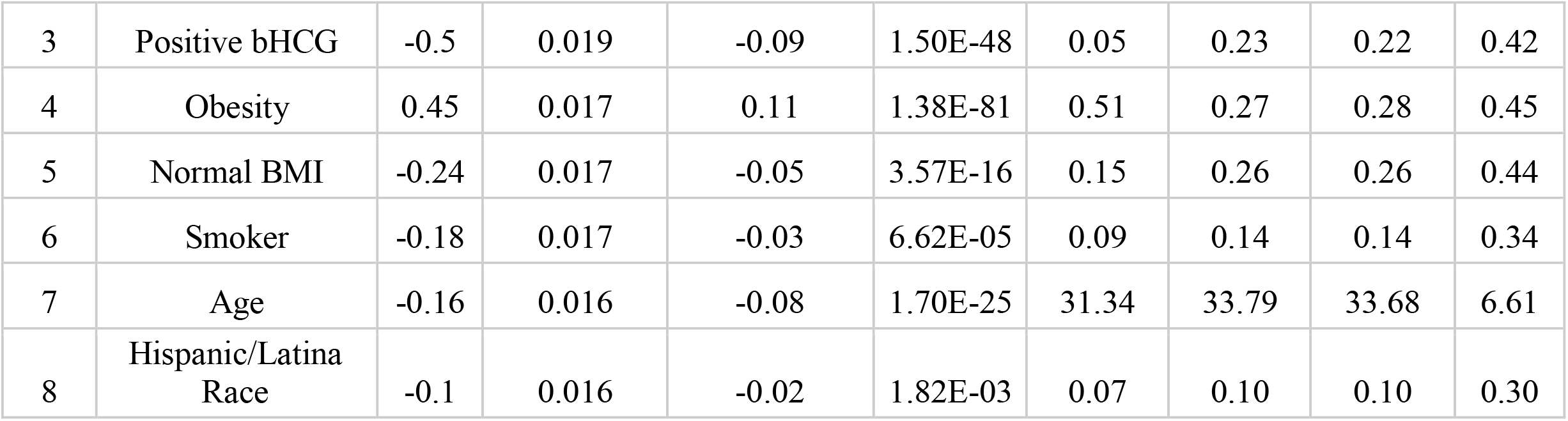
Most significant variables for PCOS diagnosis prediction in Model I.

**Table 3.** Most significant variables for PCOS diagnosis prediction in Model II.

| Rank | Variables | $\beta$ | $\beta$ - %95 CI | Y-correlation | p-value | Y1-mean | Y0-mean | All-mean | All-std |
| --- | --- | --- | --- | --- | --- | --- | --- | --- | --- |
| 0 | MLP Score | 0.61 | 0.023 | 0.26 | 2.13E-142 | 0.12 | 0.04 | 0.04 | 0.06 |
| 1 | Intercept | -0.44 | - | - | - | - | - | - | - |
| 2 | Age | -0.27 | 0.015 | -0.08 | 2.26E-31 | 31.01 | 33.79 | 33.69 | 6.61 |
| 3 | Gravidity | -0.26 | 0.016 | -0.09 | 2.35E-63 | 1.42 | 2.08 | 2.06 | 1.39 |
| 4 | Obesity | 0.21 | 0.016 | 0.03 | 9.60E-06 | 0.34 | 0.27 | 0.27 | 0.44 |
| 5 | Positive bHCG | -0.19 | 0.017 | -0.06 | 4.14E-21 | 0.10 | 0.23 | 0.23 | 0.42 |
| 6 | Hispanic/Latina Race | -0.18 | 0.016 | -0.02 | 2.69E-03 | 0.06 | 0.10 | 0.10 | 0.30 |
| 7 | Normal BP | 0.16 | 0.015 | 0.03 | 1.37E-07 | 0.60 | 0.51 | 0.51 | 0.50 |
| 8 | Normal BMI | 0.15 | 0.016 | 0.03 | 8.57E-07 | 0.34 | 0.26 | 0.26 | 0.44 |
| 9 | Negative bHCG | 0.12 | 0.015 | 0.06 | 1.44E-22 | 0.37 | 0.23 | 0.23 | 0.42 |
| 10 | HDL | 0.08 | 0.015 | 0.01 | 1.03E-10 | 52.13 | 51.59 | 51.61 | 7.86 |

**Table 4.** Most significant variables for PCOS diagnosis prediction in Model III.

| Rank | Variables | $\beta$ | $\beta$ - %95 CI | Y-correlation | p-value | Y1-mean | Y0-mean | All-mean | All-std |
| --- | --- | --- | --- | --- | --- | --- | --- | --- | --- |
| 0 | MLP Score | 0.6 | 0.023 | 0.26 | 7.41E-142 | 0.10 | 0.04 | 0.04 | 0.05 |
| 1 | Intercept | - | - | - | - | - | - | - | - |
| 2 | Age | -0.25 | 0.015 | -0.08 | 5.91E-30 | 31.16 | 33.79 | 33.69 | 6.61 |
| 3 | Gravidity | -0.24 | 0.016 | -0.09 | 2.47E-63 | 1.46 | 2.08 | 2.06 | 1.39 |
| 4 | Positive bHCG | -0.20 | 0.017 | -0.06 | 3.59E-20 | 0.11 | 0.23 | 0.23 | 0.42 |
| 5 | Obesity | 0.19 | 0.016 | 0.03 | 2.73E-06 | 0.34 | 0.27 | 0.27 | 0.44 |
| 6 | Normal BP | 0.17 | 0.015 | 0.04 | 3.94E-08 | 0.60 | 0.51 | 0.51 | 0.50 |
| 7 | Hispanic/Latina Race | -0.15 | 0.016 | -0.02 | 2.00E-03 | 0.06 | 0.10 | 0.10 | 0.30 |
| 8 | Normal BMI | 0.14 | 0.016 | 0.03 | 6.76E-06 | 0.33 | 0.26 | 0.26 | 0.44 |
| 9 | Black/African American Race | 0.13 | 0.015 | 0.02 | 2.03E-03 | 0.46 | 0.40 | 0.41 | 0.49 |
| 10 | Negative bHCG | 0.12 | 0.015 | 0.06 | 2.20E-25 | 0.37 | 0.23 | 0.23 | 0.42 |
| 11 | HDL | 0.09 | 0.015 | 0.01 | 4.06E-12 | 52.04 | 51.59 | 51.61 | 7.86 |

**Table 5.** Most significant variables for PCOS diagnosis prediction in Model IV.

| Rank | Variables | $\beta$ | $\beta$ - %95 CI | Y-correlation | p-value | Y1-mean | Y0-mean | All-mean | All-std |
| --- | --- | --- | --- | --- | --- | --- | --- | --- | --- |
| 0 | MLP Score | 0.7 | 0.024 | 0.36 | 0.00E-01 | 0.20 | 0.07 | 0.08 | 0.10 |
| 1 | Intercept | -0.44 | - | - | - | - | - | - | - |
| 2 | Gravidity | -0.37 | 0.017 | -0.14 | 2.17E-135 | 1.36 | 2.08 | 2.02 | 1.39 |
| 3 | Positive bHCG | -0.34 | 0.017 | -0.10 | 2.23E-65 | 0.08 | 0.23 | 0.22 | 0.41 |
| 4 | Obesity | 0.31 | 0.015 | 0.10 | 2.86E-66 | 0.43 | 0.27 | 0.28 | 0.45 |
| 5 | Age | -0.21 | 0.015 | -0.10 | 1.91E-52 | 31.26 | 33.79 | 33.59 | 6.62 |
| 6 | Hispanic/Latina Race | -0.12 | 0.015 | -0.03 | 2.34E-06 | 0.07 | 0.10 | 0.10 | 0.29 |
| 7 | Smoker | -0.08 | 0.015 | -0.02 | 3.00E-04 | 0.11 | 0.14 | 0.14 | 0.34 |
| 8 | Hypertension | 0.07 | 0.015 | 0.04 | 3.63E-12 | 0.28 | 0.21 | 0.22 | 0.41 |
| 9 | Education – Some College/Technical/ Vocational School | 0.06 | 0.014 | 0.03 | 1.55E-04 | 0.18 | 0.15 | 0.15 | 0.36 |
| 10 | Negative bHCG | -0.05 | 0.015 | 0.05 | 2.29E-16 | 0.31 | 0.23 | 0.24 | 0.42 |

For Model I, the parsimonious predictive model achieved an AUC (SD) of 82.3% (1.7). The MLP score (β = 0.71) and obesity (β = 0.45) were positively correlated with PCOS diagnosis. Pregnancy (gravidity β = -0.53; positive pregnancy test β = -0.50), normal BMI (β = -0.24), smoking (β = -0.18), age (β = -0.16), and Hispanic race (β = -0.10) were inversely correlated with PCOS diagnosis as shown in Table 2.

For Model II, the parsimonious predictive model achieved an AUC (SD) of 77.6% (1.3). The MLP score (β = 0.61), obesity (β = 0.21), normal BMI (β = 0.15), normal blood pressure (β = 0.16), negative pregnancy test (β = 0.12), and normal HDL (β = 0.08) were positively correlated with undiagnosed PCOS. Age (β = -0.27), pregnancy (gravidity β = -0.26; positive pregnancy test β = -0.19), and Hispanic race (β = -0.18) were inversely correlated with undiagnosed PCOS as show in Table 3.

For Model III, the parsimonious predictive model achieved an AUC (SD) of 77.4% (1.6). The MLP score (β = 0.60), obesity (β = 0.19), normal blood pressure (β = 0.17), normal BMI (β = 0.14), Black race (0.13), negative pregnancy test (β = 0.12), and normal HDL (β = 0.09) were positively correlated with undiagnosed PCOS. Age (β = -0.25), pregnancy (gravidity β = -0.24; positive pregnancy test β = - 0.20), and Hispanic race (β = -0.15) were inversely correlated with undiagnosed PCOS as show in Table 4.

For Model IV, the parsimonious predictive model achieved an AUC (SD) of 79.1% (1.1). The MLP score (β = 0.7), obesity (β = 0.31), normal BMI (β = 0.15), hypertension (β = 0.07) and some higher degree of education, such as college or vocational/technical school (β = 0.06) were positively correlated with PCOS diagnosis. Age (β = -0.21), pregnancy (gravidity β = -0.37; positive pregnancy test β = -0.34; negative pregnancy test β = -0.05), Hispanic race (β = -0.12), and smoking (β = -0.08) were inversely correlated with PCOS diagnosis as shown in Table 5.

GBT models had the highest performance. Predictions of PCOS in a test set of patients not used during algorithm training achieved 85%, 81%, 80%, and 82% AUC for Models I, II, III, and IV, respectively. We also report the performance with the logistic regression model (LR-L1) after SFS and the performance when using our developed MLP score alongside variables selected via recursive feature elimination (LR-L2-MLP score). Supplementary Table 6 displays features for each model, associated with LR-L1 algorithm after SFS. As we hypothesized, developing models using the MLP score (LR-L2-MLP score) leads to improvement of the performance of linear models (LR-L1) for Models I, II, III, and IV, respectively from 79%, 72%, 73%, and 75% AUC to 82%, 78%, 77%, and 79% AUC. Table 6 details the models with the best performance (highest AUC) using all 51 features before and after statistical feature selection (SFS). In Table 6, the means and standard deviations of AUC and weighted-F1 scores on the test set over the five repetitions are listed. Supplementary Table 7 displays the performance of all models and all algorithms, before and after statistical feature selection (SFS).

**Table 6.** Model performance over the test set, in the format of mean percentage (SD percentage) over 5 repetitions.

|  | Model I |  | Model II |  | Model III |  | Model IV |  |
| --- | --- | --- | --- | --- | --- | --- | --- | --- |
|  | AUC | F1-weighted | AUC | F1-weighted | AUC | F1-weighted | AUC | F1-weighted |
| Best full models before SFS | XGBoost (51 features) |  | XGBoost (51 features) |  | XGBoost (51 features) |  | XGBoost (51 features) |  |
|  | 85.2 (1.8) | 94.5 (0.2) | 80.6 (0.5) | 95.1 (0.2) | 80.4 (0.7) | 94.8 (0.1) | 81.8 (1.4) | 91.1 (0.4) |
| Best full models after SFS | XGBoost (14 features) |  | XGBoost (16 features) |  | XGBoost (17 features) |  | XGBoost (17 features) |  |
|  | 83.6 (1.7) | 94.5 (0.2) | 80.5 (0.7) | 95.1 (0.2) | 79.8 (1.1) | 94.8 (0.1) | 81.1 (1.3) | 90.9 (0.3) |
|  | LR-L1 (14 features) |  | LR-L1 (16 features) |  | LR-L1 (17 features) |  | LR-L1 (17 features) |  |
|  | 79.2 (1.9) | 93.9 (0.2) | 71.7 (0.9) | 94.7 (0.1) | 72.9 (2.1) | 94.4 (0.1) | 74.8 (1.1) | 89.7 (0.3) |
|  | Parsimonious models<br>LR-L2-MLP score (8 features) |  | Parsimonious models<br>LR-L2-MLP score (10 features) |  | Parsimonious models<br>LR-L2-MLP score (11 features) |  | Parsimonious models<br>LR-L2-MLP score (10 features) |  |
|  | 82.3 (1.7) | 94.5 (0.1) | 77.6 (1.3) | 95.1 (0.1) | 77.4 (1.6) | 94.9 (0.1) | 79.1 (1.1) | 90.8 (0.3) |

## Discussion

Evaluating an at-risk population for PCOS is essential for early diagnosis and initiating multi-disciplinary care with the goal of reducing health risks (endometrial hyperplasia/cancer), infertility and pregnancy complications, and chronic disease burden including cardiometabolic disorders associated with PCOS. Retrospective analysis of the at-risk population within an urban health center allows for assessment of factors predictive of diagnosis. Of note, the study sample represents a population of patients who had any visit to BMC for primary care, obstetrics and gynecology, endocrinology, family medicine, or general internal medicine and does not represent a random sample. While this is not a population level assessment, our model is applicable to patients with high suspicion for PCOS who interact with the healthcare system.

The ranked list of variables, from the most predictive to the least predictive of the PCOS outcome, informed the main drivers of the predictive models. For example, non-gravidity, high levels of LH, low levels of FSH, obesity, and higher BMI increase the likelihood of PCOS. These variables are consistent with key variables from other models and in the pathophysiology of PCOS. The overall predictive accuracy was high for all models, suggesting that a predictive model may assist in early detection of PCOS within those at risk in an electronically interfaced medical record. Furthermore, we found that non-linear models had superior predictive capacity compared to linear models for all four model outcomes, potentially allowing for inclusion of non-linear reproductive hormone relationships.

When assessing patients who received a diagnosis of PCOS (Model I), the most predictive factors related to diagnosis were hormone levels (as captured by the MLP score) and obesity, a clinical factor in supporting a PCOS diagnosis. Specifically, there is a non-linear relationship between reproductive hormones such as FSH, LH, and estradiol. Often these hormonal lab tests are obtained randomly in those with oligomenorrhea, and it is also common to find an elevated FSH to LH ratio. A concern may also be the misclassification of hypothalamic amenorrhea into the group classified as PCOS where the FSH and LH levels would be low or suppressed, or in the setting of premature ovarian insufficiency, notable by an elevated FSH and low estradiol. The MLP score allows for the diversity of relationships of these hormone levels and was trained using a neural network to appropriately classify PCOS. Additionally, prior pregnancy (gravidity) and a positive pregnancy test were negatively associated with a diagnosis of PCOS, consistent with the underlying increased risk of infertility due to oligo-ovulation. Normal BMI and smoking, a known ovarian toxicant, were negatively associated with the presence of a PCOS diagnosis, which may indicate patient characteristics that increase risk of a delayed PCOS diagnosis. These identified variables demonstrate the robustness of the model towards predicting phenotypic traits of patients with PCOS, which is aligned with the performance accuracy. While the significant factors such as hormone levels, gravidity, bHCG, and obesity identified in the model are already known to be associated with PCOS, the true impact of our model lies within the implementation of such a tool within the EHR. For example, a real-world application of this model in the clinical setting would entail integration of our model into the electronic health record system that would provide the probability of PCOS diagnosis or set a threshold for suspicion for each patient to aid a provider’s evaluation. This would lead to more timely diagnosis and optimize referrals for downstream follow-up for known clinical sequelae associated with PCOS.

When assessing patients who met diagnostic criteria without the ICD-9 label of PCOS (Models II and III), predictive factors both supported the underlying PCOS diagnosis and alluded towards factors that may contribute to missing the diagnosis despite meeting Rotterdam criteria. Similar to Model I, gravidity and a positive pregnancy test were negatively associated with Models II and III diagnosis, while obesity was positively associated with Models II and III diagnosis, consistent with Model I. Interestingly, distinct positive predictors among Models II and III were normal BMI, normal blood pressure, and normal HDL. These patients may present as the “lean” phenotype of PCOS or those with mild features, leading to underdiagnosis of PCOS. Diagnosing “lean” PCOS can be more nuanced, potentially delaying diagnosis or requiring more specialized consultation (28). Within our cohort, 1,116 individuals were identified by the model without the ICD-9 code that met Rotterdam PCOS diagnostic criteria (Model III), suggesting the predictive value of our models to identify at risk groups within a large health system and reduce delays in diagnosis. Given that women often wait over two years and see numerous health professionals before receiving a diagnosis of PCOS, the integration of high-quality AI-based diagnostic tools with the EHR could significantly contribute to more timely diagnosis (8).

Consistent with Models I, II, and III, positive pregnancy test and gravidity were both negatively associated with PCOS diagnosis in Model IV while obesity and presence of hypertension were both positively associated with the Model IV combined PCOS outcome. Some higher degree of education, such as college or vocational/technical school, was also positively associated with the outcomes of undiagnosed PCOS and combined PCOS (Models II, III, and IV), which may suggest that education status and patient’s self-advocacy for seeking care within a medical system may be implicated specifically in under-diagnosed individuals. Of note, we dropped insurance status after finding that the null was a strong predictor of PCOS, though it is interesting to note that 83% of 331 patients in this dataset with missing insurance have PCOS. Insurance status alludes to socioeconomic barriers such as access to care, which can result in a delay in timely diagnosis through either inability to seek evaluation or follow through with testing. While the implications of insurance status and social determinants of health are beyond the scope of this paper, it is important to note that persistence in seeking treatment within a fractionated health care system can be challenging financially and psychologically, as patients may need multiple evaluation or specialist’s consultation to reach the right diagnosis.

A recent systematic review investigated the utility of artificial intelligence and machine learning in the diagnosis or classification of PCOS (29). Their search ultimately included 31 studies with sample sizes ranging from 9 to 2,000 patients with PCOS. Methods employed by these models included support vector machine, K-nearest neighbor, regression models, random forest, and neural networks. Only 19% of included studies performed all major steps of training, testing, and validating their model. Furthermore, only 32% of included studies used standardized diagnostic criteria such as the Rotterdam criteria or NIH criteria. The authors found that the ROC of included studies ranged from 73-100%. Only one study sourced their data from electronic health records to build their model (30). Despite the lack of standardized model training and diagnostic criteria used in these studies, the review concluded that artificial intelligence and machine learning provide promise in detecting PCOS, allowing for an avenue for early diagnosis.

Outside of the machine learning models included in the systematic review, other predictive models have been created for earlier detection of PCOS as well as for predicting long-term health outcomes among women with a diagnosis of PCOS. One such model was created from 11,720 ovarian stimulation cycles at Peking University Third Hospital. The model used serum antimullerian hormone (AMH) and androstenedione levels, BMI, and menstrual cycle length to predict a diagnosis of PCOS. The algorithm was then developed into an online platform that is able to calculate one’s risk of PCOS given certain indicators that are inputted into the model, allowing for better screening abilities in the clinic (31). Another study created a similar model, taking into account AMH and BMI to predict a diagnosis of PCOS or other ovulatory dysfunction disorders among 2,322 women (10). They found that in women with higher BMIs and lower AMH levels could be used to predict PCOS compared to normal-weight or underweight women. Deshmukh et al. created a simple four-variable model which included free androgen index (FAI), 17-hydroxyprogesterone, AMH, and waist circumference for predicting risk of PCOS in a cross-sectional study involving 111 women with PCOS and 67 women without PCOS (32). Lastly, Joo et al. used polygenic and phenotypic risk scores to develop a PCOS risk prediction algorithm (33). They found high degrees of association between PCOS and various metabolic and endocrine disorders including obesity, type 2 diabetes, hypercholesterolemia, disorders of lipid metabolism, hypertension, and sleep apnea (33).

In addition to the goal of improved screening for PCOS, models have been created to predict long-term clinical outcomes in women with PCOS, such as ovulation, conception, and live birth (11,12). Given the increased risk of insulin resistance in women with PCOS, Gennarelli et al. created a mathematical model to predict insulin sensitivity based on variables such as BMI, waist and hip circumferences, truncal-abdominal skin folds, and serum concentrations of androgens, SHBG, triglycerides, and cholesterol (13). Models to predict non-alcoholic fatty liver disease risk among young adults with PCOS have also been generated (34). Combining earlier detection with more accurate risk stratification of clinical sequalae through predictive modeling can significantly improve the long-term health outcomes of women with PCOS. Application of our models to predict other downstream health risks after the diagnosis of PCOS is a future area of research.

Beyond the long-term health impacts of PCOS, the condition also carries a significant economic cost for our healthcare system. A study by Riestenberg et al (2022) recently estimated the total economic burden of PCOS, as well as the cost specifically for pregnancy-related complications and long-term health morbidities (2). The authors estimated the annual economic burden of PCOS to be $8 billion as of 2020 in the United States. Furthermore, the excess cost of pregnancy-related comorbidities such as gestational hypertension, gestational diabetes, and preeclampsia attributable to PCOS totals $375 million USD annually. Outside of pregnancy, the cost of long-term comorbidities associated with PCOS including stroke and type 2 diabetes mellitus was estimated at $3.9 billion USD. Meanwhile, the cost for diagnostic evaluation of PCOS was less than 2% of the total economic burden. This estimated financial burden suggests that predictive models aiding earlier diagnosis could not only reduce long-term health consequences of PCOS but also alleviate significant healthcare costs associated with the condition.

Given the high prevalence, significant healthcare burden, and heterogeneity in clinical presentation of PCOS, AI-based tools are well suited for earlier diagnosis of PCOS. Our study had many strengths. First, our machine learning models, which were highly accurate and robust in PCOS diagnosis prediction, were created using the largest sample size to date (29). Second, our model was tested and trained on a diverse Safety-Net hospital-sourced population not restricted to the context of fertility care. Third, it is the only model that incorporated three data streams (ICD-9 codes, clinical laboratory findings, and radiologic findings) and an MLP score. Fourth, the parsimonious and interpretable models were very close in achieving full model predictive accuracy, performing relatively closely to the best-performing non-linear models. Essentially, our parsimonious models “isolate” nonlinearities in hormone levels (captured by the MLP score) and linearly combine that score with other variables. Most models evaluate reproductive hormones (FSH, estradiol, LH, and SHBG) as individual variables within linear models, which does not account for the high inter- and intra-patient variability. By using non-linear mapping of the hormone values, we were able to generate a composite variable allowing for a linear function that correlates with the likelihood of an accurate prediction. Last, our variables are easily accessible in an electronic health dataset, rendering the models helpful for clinical prediction. Our study did not evaluate AMH as a predictive variable because it was not widely utilized during the time window of this data extraction corresponding with ICD-9 codes.

Despite these strengths, our model is not without limitations. First, it is only directly applicable to those who interact with the medical system and those deemed “at-risk” for a PCOS diagnosis, which would not facilitate population-based prediction. Additional studies need to be conducted in other patient populations or unselected community-based populations to validate the use of these models, especially expanding to the entire population within a health system to evaluate the accuracy of our models (35).

Second, we must interpret our data within the limitations of informative presence in EHR data. Informative presence is defined as data that is present and informed with respect to the health outcome, in this case PCOS, as well as behavioral patterns of interaction with healthcare institutions which may be additionally impacted by marginalization (36). This is an important consideration for interpreting predictive models using EHR data (36,37). Nevertheless, we were able to extract over 1000 patients who were undiagnosed with PCOS among the population, suggesting the predictive value of the modelling in identifying diagnosis gaps among specific populations within a large health system. Third, it is possible that additional examination of the medical record beyond ICD-9 diagnosis may allow for more clarification of risk in the presumed PCOS group. Last, our exclusion of concurrent endocrinopathies was chosen to avoid incorrectly including ovulation disorders caused by other endocrinopathies, but it is possible that this was an overly strict exclusion criterion.

In conclusion, this novel machine learning algorithm incorporates three data streams from a large EHR dataset to assess PCOS risk. This model can be integrated into the EHR to aid clinicians in earlier diagnosis of PCOS and connect patients to interventions and healthcare providers across their reproductive lifespan with the goal of health optimization and risk reduction.

## Acknowledgements

We would like to acknowledge Linda Rosen, the research manager of the Clinical Data Warehouse at the Boston Medical Center for procuring the dataset, and Alexis Veiga, the research assistant who provided administrative support for this project.

## Author contributions

ZZ performed the analysis and co-wrote the manuscript with a focus on the methods. VJ interpreted the findings and drafted the initial manuscript. AW conducted a literature review, contributed to interpretation of data, writing, and editing the manuscript. TW initiated the analytical approach to the research question. JC curated the initial dataset and reviewed the analysis and manuscript drafts. SM and ICP designed the study, oversaw analysis, interpretation of the findings, and manuscript drafting and revision process. All authors met ICJME criteria for authorship.

## Data availability

All datasets generated during and/or analyzed during the current study are not publicly available but are available from the corresponding author on reasonable request.

## Competing interests

The authors declare no competing interests.

## Notes

**Study funding/competing interest(s)**: This study was partially supported by National Science Foundation grants CCF-2200052, IIS-1914792, and DMS-1664644, by the NIH under grants R01 GM135930 and UL54 TR004130, and by the Boston University Kilachand Fund for Integrated Life Science and Engineering

**Disclosure Summary**: The authors declare no conflict of interest and nothing to disclose.

### Competing Interest Statement

The authors have declared no competing interest.

### Funding Statement

This study was funded by National Institutes of Health (R01 GM135930), National Institutes of Health (UL54 TR004130), Boston University Kilachand Fund for Integrated Life Science and Engineering, National Science Foundation (CCF-2200052), National Science Foundation (IIS-1914792), and National Science Foundation (DMS-1664644).

### Author Declarations

Institutional Review Board of Boston University School of Medicine and the Harvard T.H. Chan School of Public Health (Protocol # H35708) agave ethical approval for this work

### Summary of Updates

Introduction expanded for model used; vastly added to discussion to expanded on predictive model

