## Supplemental Tables for "Predicting polycystic ovary syndrome (PCOS) with machine learning algorithms from electronic health records"

Supplementary Table 1. Exclusion ICD-9 Coding Associations

| Endocrinopathies | ICD-9 Codes |
| --- | --- |
| Goiter, specified as simple | 240 |
| Goiter, unspecified | 240.9 |
| Nontoxic uninodular goiter | 241 |
| Nontoxic multinodular goiter | 241.1 |
| Thyrotoxicosis with or without goiter | 242 |
| Congenital hypothyroidism | 243 |
| Acquired hypothyroidism | 244 |
| Thyroiditis | 245 |
| Other disorders of thyroid | 246 |
| Cushing’s syndrome | 255 |
| Hyperaldosteronism | 255.1 |
| Adrenogenital disorders | 255.2 |
| Other corticoadrenal overactivity | 255.3 |
| Corticoadrenal insufficiency | 255.4 |
| Other adrenal hypofunction | 255.5 |
| Medulloadrenal hyperfunction | 255.6 |
| Other specified disorders of adrenal glands | 255.8 |
| Unspecified disorder of adrenal glands | 255.9 |
| Other ovarian dysfunction | 256.8 |
| Malignancy | ICD-9 Codes |
| Malignant neoplasm of corpus uteri, except isthmus | 182 |

Supplementary Table 2. PCOS models generated from the Boston Medical Center Clinical Data Warehouse.

| PCOS (ICD-9 256.4) | IM | HA | PCOM | Model I | Model II | Model III | Model IV | Count | Percent (%) |
| --- | --- | --- | --- | --- | --- | --- | --- | --- | --- |
| 0 | 0 | 0 | 0 | 0 | 0 | 0 | 0 | 17,834 | 58.3 |
| 1 | 0 | 0 | 0 | 1 | exclude | exclude | 1 | 313 | 1 |
| 0 | 1 | 0 | 0 | 0 | 0 | 0 | 0 | 5,012 | 16.4 |
| 0 | 0 | 1 | 0 | 0 | 0 | 0 | 0 | 5,035 | 16.5 |
| 0 | 0 | 0 | 1 | 0 | 0 | 0 | 0 | 275 | 0.8 |
| 1 | 1 | 0 | 0 | 1 | exclude | exclude | 1 | 248 | 0.8 |
| 1 | 0 | 1 | 0 | 1 | exclude | exclude | 1 | 376 | 1.2 |
| 1 | 0 | 0 | 1 | 1 | exclude | exclude | 1 | 5 | 0 |
| 0 | 1 | 1 | 0 | exclude | 1 | 1 | 1 | 1,037 | 3.4 |
| 0 | 1 | 0 | 1 | exclude | exclude | 1 | 1 | 35 | 0.1 |
| 0 | 0 | 1 | 1 | exclude | exclude | 1 | 1 | 25 | 0.1 |
| 1 | 1 | 1 | 0 | 1 | exclude | exclude | 1 | 368 | 1.2 |
| 1 | 1 | 0 | 1 | 1 | exclude | exclude | 1 | 4 | 0 |
| 1 | 0 | 1 | 1 | 1 | exclude | exclude | 1 | 9 | 0 |
| 0 | 1 | 1 | 1 | exclude | 1 | 1 | 1 | 19 | 0.1 |
| 1 | 1 | 1 | 1 | 1 | exclude | exclude | 1 | 6 | 0 |
| Total Patients |  |  |  | 29,485 | 29,212 | 29,272 | 30,601 | 30,601 | 100 |

Supplementary Table 3. Summary of missingness in continuous variables.

|  | Model I | | Model II | | Model III | | Model IV | |
| --- | --- | --- | --- | --- | --- | --- | --- | --- |
| variable | # of missing | Imputed value | # of missing | Imputed value | # of missing | Imputed value | # of missing | Imputed value |
| Testosterone | 27365 | 29.8 | 27201 | 29.0 | 27251 | 29.0 | 28115 | 32.0 |
| Free  Testosterone | 26975 | 3.0 | 26843 | 3.0 | 26887 | 3.0 | 27682 | 3.3 |
| Bioavailable Testosterone | 26987 | 6.05 | 26857 | 6.2 | 26901 | 6.0 | 27698 | 6.7 |
| Gravidity | 11282 | 2.0 | 11141 | 2.0 | 11148 | 2.0 | 11537 | 2.0 |
| Age at Menarche | 13120 | 12.0 | 12984 | 12.0 | 12996 | 12.0 | 13430 | 12.0 |
| Total Cholesterol | 17905 | 172.0 | 17688 | 172.0 | 17710 | 172.0 | 18350 | 172.0 |
| HDL | 18104 | 51.0 | 17887 | 51.0 | 17909 | 51.0 | 18562 | 51.0 |
| LDL | 18955 | 101.0 | 18746 | 101.0 | 18772 | 101.0 | 19508 | 101.0 |
| TSH | 20604 | 1.22 | 20303 | 1.21 | 20339 | 1.21 | 21296 | 1.2 |
| A1C | 23139 | 5.4 | 22949 | 5.4 | 22995 | 5.4 | 23954 | 5.4 |
| FSH | 24338 | 5.0 | 24205 | 5.1 | 24236 | 5.1 | 24893 | 5.0 |
| LH | 25941 | 6.3 | 25823 | 6.1 | 25862 | 6.1 | 26641 | 6.5 |
| SHBG | 27181 | 39.0 | 27049 | 41.0 | 27094 | 41.0 | 27942 | 38.0 |
| Estradiol | 27331 | 58.0 | 27072 | 58.0 | 27120 | 58.0 | 28235 | 58.0 |

Supplementary Table 4. Representative patient statistics.

|  | Model I | | | Model II | | | Model III | | | Model IV | | |
| --- | --- | --- | --- | --- | --- | --- | --- | --- | --- | --- | --- | --- |
| Variable | Y1-mean | Y0-mean | p-value | Y1-mean | Y0-mean | p-value | Y1-mean | Y0-mean | p-value | Y1-mean | Y0-mean | p-value |
| Gravidity | 1.28 | 2.08 | 4.55E-78 | 1.42 | 2.08 | 2.35E-63 | 1.46 | 2.08 | 2.47E-63 | 1.36 | 2.08 | 2.17E-135 |
| LH | 8.03 | 6.53 | 3.65E-69 | 7.72 | 6.35 | 5.07E-35 | 7.69 | 6.35 | 2.81E-36 | 8.04 | 6.70 | 4.47E-94 |
| SHBG | 38.48 | 39.90 | 1.87E-58 | 40.03 | 41.75 | 4.89E-32 | 40.01 | 41.75 | 1.87E-32 | 37.84 | 38.94 | 8.51E-78 |
| FSH | 4.94 | 5.24 | 9.46E-44 | 5.28 | 5.35 | 3.04E-30 | 5.26 | 5.34 | 1.42E-32 | 5.04 | 5.22 | 1.55E-68 |
| Obesity | 0.51 | 0.27 | 1.38E-81 | 0.34 | 0.27 | 9.60E-06 | 0.34 | 0.27 | 2.73E-06 | 0.43 | 0.27 | 2.86E-66 |
| Positive bHCG | 0.05 | 0.23 | 1.50E-48 | 0.10 | 0.23 | 4.14E-21 | 0.11 | 0.23 | 3.59E-20 | 0.08 | 0.23 | 2.23E-65 |
| Age | 31.34 | 33.79 | 1.70E-25 | 31.01 | 33.79 | 2.26E-31 | 31.16 | 33.79 | 5.91E-30 | 31.26 | 33.79 | 1.91E-52 |
| Obese BMI | 0.45 | 0.25 | 7.99E-57 | 0.30 | 0.25 | 1.08E-03 | 0.30 | 0.25 | 7.69E-04 | 0.38 | 0.25 | 1.03E-44 |
| HDL | 50.21 | 51.58 | 1.15E-14 | 52.13 | 51.59 | 1.03E-10 | 52.04 | 51.59 | 4.06E-12 | 51.04 | 51.58 | 3.65E-25 |
| Negative bHCG | 0.26 | 0.23 | 1.99E-01 | 0.37 | 0.23 | 1.44E-22 | 0.37 | 0.23 | 2.20E-25 | 0.31 | 0.23 | 2.29E-16 |
| Total Cholesterol | 174.77 | 173.12 | 1.70E-06 | 173.34 | 173.12 | 2.82E-10 | 173.30 | 173.12 | 7.72E-11 | 174.10 | 173.12 | 1.48E-15 |
| Hypertension | 0.31 | 0.21 | 6.02E-14 | 0.25 | 0.21 | 7.25E-02 | 0.25 | 0.21 | 8.32E-02 | 0.28 | 0.21 | 3.63E-12 |
| LDL | 102.51 | 101.51 | 2.10E-03 | 101.28 | 101.51 | 1.66E-06 | 101.37 | 101.51 | 6.31E-07 | 101.99 | 101.51 | 4.06E-07 |
| Hispanic/Latina Race | 0.07 | 0.10 | 1.82E-03 | 0.06 | 0.10 | 2.69E-03 | 0.06 | 0.10 | 2.00E-03 | 0.07 | 0.10 | 2.34E-06 |
| Estradiol | 60.70 | 59.37 | 7.32E-03 | 61.40 | 59.39 | 9.91E-04 | 61.58 | 59.39 | 3.78E-04 | 61.11 | 59.38 | 3.49E-06 |
| Education – Some College/Technical/ Vocational School | 0.18 | 0.15 | 3.44E-02 | 0.19 | 0.15 | 3.32E-03 | 0.19 | 0.15 | 4.65E-03 | 0.18 | 0.15 | 1.55E-04 |
| Smoker | 0.09 | 0.14 | 6.62E-05 | 0.12 | 0.14 | 4.11E-01 | 0.12 | 0.14 | 5.73E-01 | 0.11 | 0.14 | 3.00E-04 |
| TSH | 1.31 | 1.26 | 5.24E-02 | 1.28 | 1.26 | 8.51E-03 | 1.28 | 1.26 | 3.51E-03 | 1.30 | 1.26 | 8.49E-03 |
| Elevated BP | 0.12 | 0.10 | 2.85E-02 | 0.11 | 0.10 | 4.29E-01 | 0.11 | 0.10 | 3.35E-01 | 0.12 | 0.10 | 9.50E-03 |
| Marital Status: Single | 0.77 | 0.76 | 6.82E-01 | 0.81 | 0.76 | 1.31E-03 | 0.81 | 0.76 | 2.78E-03 | 0.79 | 0.76 | 9.87E-03 |
| Gastric Bypass History | 0.00 | 0.01 | 6.50E-02 | 0.00 | 0.01 | 3.31E-01 | 0.00 | 0.01 | 2.68E-01 | 0.00 | 0.01 | 1.25E-02 |
| Age at Menarche | 12.10 | 12.23 | 8.95E-02 | 12.18 | 12.23 | 3.10E-01 | 12.18 | 12.23 | 1.88E-01 | 12.13 | 12.23 | 1.46E-02 |
| Overweight ICD-9 278.02 | 0.04 | 0.03 | 2.58E-01 | 0.04 | 0.03 | 2.96E-01 | 0.04 | 0.03 | 1.50E-01 | 0.04 | 0.03 | 3.30E-02 |
| Normal BMI | 0.15 | 0.26 | 3.57E-16 | 0.34 | 0.26 | 8.57E-07 | 0.33 | 0.26 | 6.76E-06 | 0.23 | 0.26 | 4.80E-02 |
| A1C | 5.43 | 5.42 | 1.19E-01 | 5.42 | 5.42 | 2.45E-01 | 5.42 | 5.42 | 3.19E-01 | 5.43 | 5.42 | 5.26E-02 |
| Education: Declined | 0.03 | 0.04 | 6.85E-01 | 0.02 | 0.04 | 5.52E-02 | 0.02 | 0.04 | 5.69E-02 | 0.03 | 0.04 | 6.20E-02 |
| Marital Status: Separated | 0.01 | 0.01 | 2.11E-01 | 0.01 | 0.01 | 4.24E-01 | 0.01 | 0.01 | 4.77E-01 | 0.01 | 0.01 | 7.94E-02 |
| Black/African American Race | 0.40 | 0.40 | 1.00E+00 | 0.45 | 0.40 | 1.84E-02 | 0.46 | 0.40 | 2.03E-03 | 0.43 | 0.40 | 1.05E-01 |
| Marital Status: Married | 0.19 | 0.20 | 8.72E-01 | 0.16 | 0.20 | 3.69E-02 | 0.17 | 0.20 | 6.70E-02 | 0.18 | 0.20 | 1.30E-01 |
| Race: Other | 0.04 | 0.03 | 2.54E-01 | 0.04 | 0.03 | 4.69E-01 | 0.04 | 0.03 | 6.82E-01 | 0.04 | 0.03 | 1.64E-01 |
| Marital Status: Widowed | 0.00 | 0.00 | 6.47E-01 | 0.00 | 0.00 | 7.26E-01 | 0.00 | 0.00 | 7.08E-01 | 0.00 | 0.00 | 3.85E-01 |
| Normal BP | 0.46 | 0.51 | 1.74E-02 | 0.60 | 0.51 | 1.37E-07 | 0.60 | 0.51 | 3.94E-08 | 0.53 | 0.51 | 3.92E-01 |
| White/Caucasian Race | 0.26 | 0.27 | 9.77E-01 | 0.25 | 0.27 | 8.19E-01 | 0.25 | 0.27 | 5.44E-01 | 0.25 | 0.27 | 6.43E-01 |
| Education: high school graduate | 0.25 | 0.24 | 9.12E-01 | 0.25 | 0.24 | 8.19E-01 | 0.26 | 0.24 | 7.47E-01 | 0.25 | 0.24 | 6.63E-01 |
| Human immuno virus ICD-9 042 | 0.00 | 0.01 | 2.42E-01 | 0.01 | 0.01 | 9.97E-01 | 0.01 | 0.01 | 9.80E-01 | 0.00 | 0.01 | 7.04E-01 |
| Education: Some high school | 0.17 | 0.20 | 2.40E-01 | 0.20 | 0.20 | 9.97E-01 | 0.20 | 0.20 | 9.75E-01 | 0.19 | 0.20 | 7.09E-01 |
| Education: I did not attend school | 0.03 | 0.04 | 6.51E-01 | 0.04 | 0.04 | 9.65E-01 | 0.04 | 0.04 | 9.95E-01 | 0.04 | 0.04 | 7.48E-01 |
| Marital Status: Divorced | 0.01 | 0.01 | 1.00E+00 | 0.01 | 0.01 | 4.77E-01 | 0.01 | 0.01 | 5.34E-01 | 0.01 | 0.01 | 7.68E-01 |
| Education: Pre-registration | 0.01 | 0.01 | 2.33E-01 | 0.00 | 0.01 | 9.16E-01 | 0.00 | 0.01 | 8.72E-01 | 0.01 | 0.01 | 8.21E-01 |
| Education: 8th grade or less | 0.04 | 0.04 | 9.70E-01 | 0.04 | 0.04 | 8.63E-01 | 0.03 | 0.04 | 8.46E-01 | 0.04 | 0.04 | 8.23E-01 |
| Education: other | 0.01 | 0.01 | 9.97E-01 | 0.00 | 0.01 | 8.14E-01 | 0.00 | 0.01 | 7.51E-01 | 0.01 | 0.01 | 8.50E-01 |
| Fem Pelv Inflam Dis NOS ICD-9 614.9 | 0.00 | 0.01 | 1.60E-01 | 0.01 | 0.01 | 9.84E-01 | 0.01 | 0.01 | 6.27E-01 | 0.01 | 0.01 | 9.08E-01 |
| American Indian/Native American Race | 0.00 | 0.01 | 9.67E-01 | 0.00 | 0.01 | 9.88E-01 | 0.00 | 0.01 | 9.72E-01 | 0.00 | 0.01 | 9.24E-01 |
| Homeless Indicator No | 0.99 | 0.98 | 9.17E-01 | 0.98 | 0.98 | 1.00E+00 | 0.98 | 0.98 | 9.97E-01 | 0.99 | 0.98 | 9.33E-01 |
| Homeless Indicator Yes | 0.01 | 0.02 | 9.23E-01 | 0.02 | 0.02 | 1.00E+00 | 0.02 | 0.02 | 9.98E-01 | 0.01 | 0.02 | 9.40E-01 |
| Middle Eastern Race | 0.01 | 0.01 | 1.00E+00 | 0.01 | 0.01 | 7.67E-01 | 0.01 | 0.01 | 8.45E-01 | 0.01 | 0.01 | 9.42E-01 |
| Education: graduated college/technical school/vocational training | 0.20 | 0.21 | 9.08E-01 | 0.21 | 0.21 | 1.00E+00 | 0.21 | 0.21 | 1.00E+00 | 0.20 | 0.21 | 9.59E-01 |
| Asian Race | 0.04 | 0.05 | 7.03E-01 | 0.05 | 0.05 | 7.76E-01 | 0.05 | 0.05 | 9.39E-01 | 0.04 | 0.05 | 9.79E-01 |
| Native Hawaiian/Pacific Islander Race | 0.00 | 0.00 | 8.49E-01 | 0.00 | 0.00 | 9.78E-01 | 0.00 | 0.00 | 9.85E-01 | 0.00 | 0.00 | 9.86E-01 |
| Pure Hypercholesterolemia_ICD-9 272.0 | 0.01 | 0.01 | 1.00E+00 | 0.01 | 0.01 | 9.94E-01 | 0.01 | 0.01 | 9.74E-01 | 0.01 | 0.01 | 9.86E-01 |
| Overweight BMI | 0.18 | 0.19 | 4.58E-01 | 0.22 | 0.19 | 3.32E-01 | 0.22 | 0.19 | 1.62E-01 | 0.20 | 0.19 | 9.90E-01 |
| History of Hysterosalpingogram | 0.05 | 0.06 | 1.00E+00 | 0.04 | 0.06 | 9.86E-01 | 0.04 | 0.06 | 9.93E-01 | 0.05 | 0.06 | 1.00E+00 |
| Marital Status: Other | 0.02 | 0.02 | 8.25E-01 | 0.01 | 0.02 | 8.48E-01 | 0.01 | 0.02 | 7.48E-01 | 0.02 | 0.02 | 1.00E+00 |

Supplementary Table 5. Highly correlated variables and the retained variable.

| **Highly correlated pairs** | **The variable we kept among highly correlated pairs** |
| --- | --- |
| Marital Status – Married Marital Status – Single | Marital Status – Married |
| Homeless indicator – No  Homeless indicator – Yes | Homeless Indicator – Yes |
| Obesity Obese BMI | Obesity |
| Total Cholesterol  LDL | Total Cholesterol kept in Model I and Model IV 'LDL' kept in Model II and Model III |

Supplementary Table 6. Features for each model, associated with LR-L1 algorithm after SFS.

| Model I |  |  |  |  |  |  |  |  |
| --- | --- | --- | --- | --- | --- | --- | --- | --- |
| Rank | Variables | $\beta$ | Ycorrelation | p-value | Y1-mean | Y0-mean | All-mean | All-std |
| 0 | Intercept | -0.62 | - | - | - | - | - | - |
| 1 | FSH | -0.6 | -0.03 | 9.46E-44 | 4.94 | 5.24 | 5.23 | 2.30 |
| 2 | LH | 0.54 | 0.12 | 3.65E-69 | 8.03 | 6.53 | 6.59 | 2.70 |
| 3 | Positive bHCG | -0.54 | -0.09 | 1.50E-48 | 0.05 | 0.23 | 0.22 | 0.42 |
| 4 | Gravidity | -0.53 | -0.12 | 4.55E-78 | 1.28 | 2.08 | 2.04 | 1.39 |
| 5 | Obesity | 0.49 | 0.11 | 1.38E-81 | 0.51 | 0.27 | 0.28 | 0.45 |
| 6 | Age | -0.25 | -0.08 | 1.70E-25 | 31.34 | 33.79 | 33.68 | 6.61 |
| 7 | Normal BMI | -0.22 | -0.05 | 3.57E-16 | 0.15 | 0.26 | 0.26 | 0.44 |
| 8 | Smoker | -0.18 | -0.03 | 6.62E-05 | 0.09 | 0.14 | 0.14 | 0.34 |
| 9 | Total Cholesterol | 0.11 | 0.02 | 1.70E-06 | 174.77 | 173.12 | 173.19 | 20.85 |
| 10 | Hispanic/Latina Race | -0.11 | -0.02 | 1.82E-03 | 0.07 | 0.10 | 0.10 | 0.30 |
| 11 | Estradiol | 0.08 | 0.02 | 7.32E-03 | 60.70 | 59.37 | 59.43 | 16.54 |
| 12 | HDL | -0.08 | -0.04 | 1.15E-14 | 50.21 | 51.58 | 51.52 | 7.78 |
| 13 | Hypertension | 0.08 | 0.05 | 6.02E-14 | 0.31 | 0.21 | 0.22 | 0.41 |
| 14 | SHBG | 0.01 | -0.03 | 1.87E-58 | 38.48 | 39.90 | 39.83 | 9.94 |
| Model II |  |  |  |  |  |  |  |  |
| Rank | Variables | $\beta$_ | Y-correlation | p-value | Y1-mean | Y0-mean | All-mean | All-std |
| 0 | Age | -0.31 | -0.08 | 2.26E-31 | 31.01 | 33.79 | 33.69 | 6.61 |
| 1 | Intercept | -0.31 | - | - | - | - | - | - |
| 2 | LH | 0.29 | 0.09 | 5.07E-35 | 7.72 | 6.35 | 6.40 | 2.73 |
| 3 | Gravidity | -0.29 | -0.09 | 2.35E-63 | 1.42 | 2.08 | 2.06 | 1.39 |
| 4 | Obesity | 0.29 | 0.03 | 9.60E-06 | 0.34 | 0.27 | 0.27 | 0.44 |
| 5 | Positive bHCG | -0.2 | -0.06 | 4.14E-21 | 0.10 | 0.23 | 0.23 | 0.42 |
| 6 | Negative bHCG | 0.19 | 0.06 | 1.44E-22 | 0.37 | 0.23 | 0.23 | 0.42 |
| 7 | FSH | -0.16 | 0.00 | 3.04E-30 | 5.28 | 5.35 | 5.34 | 2.51 |
| 8 | Normal BMI | 0.15 | 0.03 | 8.57E-07 | 0.34 | 0.26 | 0.26 | 0.44 |
| 9 | Hispanic/Latina Race | -0.13 | -0.02 | 2.69E-03 | 0.06 | 0.10 | 0.10 | 0.30 |
| 10 | Normal BP | 0.13 | 0.03 | 1.37E-07 | 0.60 | 0.51 | 0.51 | 0.50 |
| 11 | SHBG | -0.08 | -0.03 | 4.89E-32 | 40.03 | 41.75 | 41.68 | 9.23 |
| 12 | HDL | 0.06 | 0.01 | 1.03E-10 | 52.13 | 51.59 | 51.61 | 7.86 |
| 13 | Education – Some College/Technical/ Vocational School | 0.06 | 0.02 | 3.32E-03 | 0.19 | 0.15 | 0.15 | 0.36 |
| 14 | Estradiol | 0.04 | 0.02 | 9.91E-04 | 61.40 | 59.39 | 59.46 | 16.82 |
| 15 | LDL | 0.01 | 0.00 | 1.66E-06 | 101.28 | 101.51 | 101.50 | 15.34 |
| 16 | TSH | 0.01 | 0.01 | 8.51E-03 | 1.28 | 1.26 | 1.26 | 0.41 |
| Model III |  |  |  |  |  |  |  |  |
| Rank | Variables | $\beta$_ | Y-correlation | p-value | Y1-mean | Y0-mean | All-mean | All-std |
| 0 | Intercept | -0.29 | - | - | - | - | - | - |
| 1 | LH | 0.29 | 0.09 | 2.81E-36 | 7.69 | 6.35 | 6.40 | 2.73 |
| 2 | Age | -0.28 | -0.08 | 5.91E-30 | 31.16 | 33.79 | 33.69 | 6.61 |
| 3 | Gravidity | -0.27 | -0.086 | 0.00 | 1.46E+00 | 2.08 | 2.06 | 1.39 |
| 4 | Obesity | 0.27 | 0.03 | 2.73E-06 | 0.34 | 0.27 | 0.27 | 0.44 |
| 5 | Negative HCG | 0.2 | 0.06 | 2.20E-25 | 0.37 | 0.23 | 0.23 | 0.42 |
| 6 | Positive HCG | -0.2 | -0.06 | 3.59E-20 | 0.11 | 0.23 | 0.23 | 0.42 |
| 7 | FSH | -0.17 | -0.01 | 1.42E-32 | 5.26 | 5.34 | 5.34 | 2.49 |
| 8 | Normal BP | 0.15 | 0.04 | 3.94E-08 | 0.60 | 0.51 | 0.51 | 0.50 |
| 9 | Normal BMI | 0.13 | 0.03 | 6.76E-06 | 0.33 | 0.26 | 0.26 | 0.44 |
| 10 | Black/African American Race | 0.12 | 0.02 | 2.03E-03 | 0.46 | 0.40 | 0.41 | 0.49 |
| 11 | Hispanic/Latina Race | -0.1 | -0.02 | 2.00E-03 | 0.06 | 0.10 | 0.10 | 0.30 |
| 12 | SHBG | -0.07 | -0.04 | 1.87E-32 | 40.01 | 41.75 | 41.68 | 9.23 |
| 13 | HDL | 0.06 | 0.01 | 4.06E-12 | 52.04 | 51.59 | 51.61 | 7.86 |
| 14 | Education – Some College/Technical/ Vocational School | 0.06 | 0.02 | 4.65E-03 | 0.19 | 0.15 | 0.15 | 0.36 |
| 15 | Estradiol | 0.05 | 0.02 | 3.78E-04 | 61.58 | 59.39 | 59.47 | 16.88 |
| 16 | TSH | 0.04 | 0.01 | 3.51E-03 | 1.28 | 1.26 | 1.26 | 0.41 |
| 17 | LDL | 0.01 | 0.00 | 6.31E-07 | 101.37 | 101.51 | 101.50 | 15.36 |
| Model IV |  |  |  |  |  |  |  |  |
| Rank | Variables | $\beta$ | Y-correlation | p-value | Y1-mean | Y0-mean | All-mean | All-std |
| 0 | LH | 0.39 | 0.13 | 4.47E-94 | 8.04 | 6.70 | 6.81 | 2.78 |
| 1 | Gravidity | -0.39 | -0.14 | 2.17E-135 | 1.36 | 2.08 | 2.02 | 1.39 |
| 2 | Obesity | 0.38 | 0.10 | 2.86E-66 | 0.43 | 0.27 | 0.28 | 0.45 |
| 3 | Intercept | -0.35 | - | - | - | - | - | - |
| 4 | Positive HCG | -0.35 | -0.10 | 2.23E-65 | 0.08 | 0.23 | 0.22 | 0.41 |
| 5 | FSH | -0.3 | -0.02 | 1.55E-68 | 5.04 | 5.22 | 5.20 | 2.09 |
| 6 | Age | -0.28 | -0.10 | 1.91E-52 | 31.26 | 33.79 | 33.59 | 6.62 |
| 7 | Hispanic/Latina Race | -0.11 | -0.03 | 2.34E-06 | 0.07 | 0.10 | 0.10 | 0.29 |
| 8 | Hypertension | 0.1 | 0.04 | 3.63E-12 | 0.28 | 0.21 | 0.22 | 0.41 |
| 9 | Smoker | -0.09 | -0.02 | 3.00E-04 | 0.11 | 0.14 | 0.14 | 0.34 |
| 10 | Estradiol | 0.06 | 0.03 | 3.49E-06 | 61.11 | 59.38 | 59.52 | 17.10 |
| 11 | Education – Some College/Technical/ Vocational School | 0.06 | 0.03 | 1.55E-04 | 0.18 | 0.15 | 0.15 | 0.36 |
| 12 | Total Cholesterol | 0.06 | 0.01 | 1.48E-15 | 174.10 | 173.12 | 173.19 | 21.00 |
| 13 | Elevated BP | 0.06 | 0.02 | 9.50E-03 | 0.12 | 0.10 | 0.10 | 0.30 |
| 14 | Negative HCG | 0.05 | 0.05 | 2.29E-16 | 0.31 | 0.23 | 0.24 | 0.42 |
| 15 | HDL | -0.01 | -0.02 | 3.65E-25 | 51.04 | 51.58 | 51.54 | 7.88 |
| 16 | SHBG | -0.01 | -0.03 | 8.51E-78 | 37.84 | 38.94 | 38.85 | 10.03 |
| 17 | TSH | 0.01 | 0.02 | 8.49E-03 | 1.30 | 1.26 | 1.27 | 0.42 |

Supplementary table 7. Performance of all models and all algorithms, before and after SFS. The means and standard deviations of AUC and weighted-F1 scores on the test set over the five repetitions are listed in the format of mean percentage (SD percentage).

|  | Model I | | Model II | | Model III | | Model IV | |
| --- | --- | --- | --- | --- | --- | --- | --- | --- |
|  | AUC | F1-weighted | AUC | F1-weighted | AUC | F1-weighted | AUC | F1-weighted |
| Models  before SFS (51 features) | LR-L1 | | LR-L1 | | LR-L1 | | LR-L1 | |
|  | 79.4 (2.0) | 93.9 (0.1) | 72.9 (0.9) | 94.8 (0.1) | 73.9 (1.9) | 94.5 (0.2) | 75.3 (1.1) | 89.8 (0.5) |
|  | SVM-L1 | | SVM-L1 | | SVM-L1 | | SVM-L1 | |
|  | 79.3 (2.0) | 93.9 (0.1) | 73.1 (0.7) | 94.7 (0.1) | 74.1 (1.9) | 94.4 (0.2) | 75.3 (1.1) | 89.7 (0.3) |
|  | XGBoost | | XGBoost | | XGBoost | | XGBoost | |
|  | 85.2 (1.8) | 94.5 (0.2) | 80.6 (0.5) | 95.1 (0.2) | 80.4 (0.7) | 94.8 (0.1) | 81.8 (1.4) | 91.1 (0.4) |
|  | RF | | RF | | RF | | RF | |
|  | 84.3 (1.7) | 94.5 (0.2) | 79.6 (0.4) | 95.1 (0.2) | 80.3 (0.9) | 94.8 (0.1) | 81.3 (1.6) | 90.9 (0.3) |
| Models  after SFS (14, 16, 17, 17 features respectively for Model I, II, III, IV) | LR-L1 | | LR-L1 | | LR-L1 | | LR-L1 | |
|  | 79.2 (1.9) | 93.9 (0.2) | 71.7 (0.9) | 94.7 (0.1) | 72.9 (2.1) | 94.4 (0.1) | 74.8 (1.1) | 89.7 (0.3) |
|  | SVM-L1 | | SVM-L1 | | SVM-L1 | | SVM-L1 | |
|  | 79.1 (1.9) | 93.8 (0.2) | 71.7 (0.9) | 94.7 (0.1) | 72.8 (2.0) | 94.4 (0.1) | 74.7 (1.0) | 89.6 (0.3) |
|  | XGBoost | | XGBoost | | XGBoost | | XGBoost | |
|  | 83.6 (1.7) | 94.5 (0.2) | 80.5 (0.7) | 95.1 (0.2) | 79.8 (1.1) | 94.8 (0.1) | 81.1 (1.3) | 90.9 (0.3) |
|  | RF | | RF | | RF | | RF | |
|  | 82.8 (2.0) | 94.5 (0.1) | 79.5 (0.5) | 95.0 (0.3) | 79.7 (0.8) | 94.9 (0.2) | 80.4 (1.3) | 90.8 (0.3) |
| Parsimonious  models LR-L2-MLP score (8, 10, 11, 10 features respectively for Model I, II, III, IV) | 82.3 (1.7) | 94.5 (0.1) | 77.6 (1.3) | 95.1 (0.1) | 77.4 (1.6) | 94.9 (0.1) | 79.1 (1.1) | 90.8 (0.3) |
